# Iron homeostasis and endometriosis risk: Genetic evidence for a shared biological link

**DOI:** 10.64898/2026.08.25.26361232

**Authors:** Veronika Denner, Christian M. Becker, Hal Drakesmith, Krina T. Zondervan, Sam Morris, Nilufer Rahmioglu

## Abstract

**STUDY QUESTION:** Is genetic liability to endometriosis associated with iron homeostasis, and is this relationship potentially causal?

**SUMMARY ANSWER:** Genetic evidence indicates that reduced systemic iron status is associated with increased risk of endometriosis, with evidence of 8 shared genome-wide significant loci and suggestive but inconsistent evidence for causal bidirectional effects.

**WHAT IS KNOWN ALREADY:** Endometriosis is a chronic inflammatory condition associated with local iron accumulation within ectopic lesions and peritoneal cavity, resulting from retrograde menstruation and altered iron homeostasis. Epidemiological studies have suggested that women with endometriosis may exhibit reduced systemic iron stores compared to women without endometriosis, reflected by lower circulating ferritin concentrations, although findings have been inconsistent and may be confounded by menstrual blood loss and inflammation. As observational studies cannot distinguish causal relationships from secondary effects or residual confounding, the potential genetic basis linking iron homeostasis and endometriosis risk remains unclear.

**STUDY DESIGN, SIZE, DURATION:** We performed genetic analyses using summary statistics from large-scale genome-wide association studies (GWAS) of endometriosis (overall and stage III/IV disease) and five iron biomarkers (serum iron, ferritin, total iron-binding capacity (TIBC), transferrin saturation, and hepcidin). Analyses included genome-wide genetic correlation using linkage disequilibrium score regression (LDSC), identification of shared genetic variants using multi-trait GWAS (MTAG) and bidirectional Mendelian randomisation to evaluate potential causal relationships.

**PARTICIPANTS/MATERIALS, SETTING, METHODS:** Iron biomarker summary statistics came from a six-cohort GWAS meta-analysis (HUNT, MGI, SardiNIA, deCODE, Interval, DBDS; N up to 257,953) of blood-derived serum iron, ferritin, transferrin saturation and TIBC (Moksnes et al., 2022). Endometriosis summary statistics came from a 24-study GWAS meta-analysis (60,674 cases, 701,926 controls; European and East Asian ancestry), 12 of which had surgically confirmed cases (Rahmioglu et al., 2023).

Genome-wide genetic correlations between iron biomarkers and endometriosis (overall and stage III/IV disease) were estimated using linkage disequilibrium score regression (LDSC), based on summary statistics aligned to the GRCh37 reference genome and restricted to HapMap3 variants. Multi-trait GWAS (MTAG) was applied to each iron biomarker jointly with endometriosis to enhance discovery of genetic loci. Shared loci were functionally annotated using reproductive and iron related tissues from GTEx v8 and blood from eQTLGen expression quantitative trait loci (eQTL) data. Bidirectional Mendelian randomisation (MR) analyses were performed using genome-wide significant variants across multiple clumping thresholds, with inverse-variance weighting (IVW) as the primary method and sensitivity analyses including weighted median, MR-Egger and MR-PRESSO.

**MAIN RESULTS AND THE ROLE OF CHANCE:** Genetic correlation analyses suggested that a genetic predisposition to endometriosis is associated with a profile of lower systemic iron availability. Specifically, genetic liability to endometriosis was associated with higher total iron-binding capacity (TIBC; rg=0.16, p=4x10^-4^), together with lower transferrin saturation (rg=-0.16, p=0.006) and lower ferritin levels (rg= -0.10, p=0.022), findings that are consistent with reduced iron stores. MTAG identified eight additional genome-wide significant loci for endometriosis and eight loci shared with iron biomarkers, including regions implicating coagulation (F5), innate immune response (IL1A), reproductive biology (WNT4), and immune and vascular pathways (e.g. ABO, STAT6). Mendelian randomisation analyses provided limited and inconsistent evidence for a causal relationship between iron status and endometriosis. Although the inverse-variance weighted (IVW) model showed nominal associations between higher ferritin levels and a lower risk of endometriosis (OR = 0.85, 95% CI 0.76–0.94; p = 0.002), and between genetic liability to endometriosis and higher TIBC (OR = 1.02, 95% CI 1.01–1.04; p = 0.006), these findings were not consistently supported by sensitivity analyses. MR-PRESSO identified a small number of pleiotropic variants, but their removal did not materially alter the results.

**LIMITATIONS, REASONS FOR CAUTION:** Iron biomarker GWAS included males and females, potentially obscuring female-specific effects. Dataset availability restricted analyses to European ancestry, limiting applicability to other populations, and to overall and stage III/IV endometriosis, precluding assessment of other disease subtypes. Heterogeneity across SNP instruments, reflected by Cochran’s Q statistics, reduced the precision of Mendelian randomisation estimates. Moreover, the genetic instruments explained only between approximately 1.0% and 18.8% of variance in the iron biomarkers, depending on the clumping threshold, which may have limited power to detect causal effects.

**WIDER IMPLICATIONS OF THE FINDINGS:** These findings suggest that endometriosis is genetically associated with reduced systemic iron availability and altered iron homeostasis. Thus, lower systemic iron status observed in women with endometriosis may not be explained solely by menstrual blood loss or dietary factors, but reflect an underlying genetic predisposition. Shared genetic loci implicate coagulation, ABO biology, and immune pathways as potential mechanisms linking iron metabolism and endometriosis. Although Mendelian randomisation did not provide consistent evidence for causality, these findings support a shared genetic architecture and warrant further investigation using female-specific GWAS, refined disease subtypes, and multi-omic approaches. Clinically, these findings suggest that low systemic iron status in women with endometriosis may reflect factors beyond established causes of iron deficiency, including an underlying genetic predisposition.

**STUDY FUNDING/COMPETING INTEREST(S):** The research was supported by a three-month post-MSc fellowship (September to December 2025), awarded to V.D. by the Nuffield Department of Population Health. N.R. is a consultant for Endogene.bio, outside this submitted work. V.D. and S.M. declare no competing interests.

**TRIAL REGISTRATION NUMBER:** N/A.

## INTRODUCTION

Endometriosis is a chronic systemic condition defined by the presence of endometrial-like tissue outside the uterus (Zondervan, Becker and Missmer, 2020). While endometriosis lesions are most commonly located in the pelvic cavity, they can also grow in more distant sites, including the thoracic cavity (Hirata, Koga and Osuga, 2020). The condition affects around ten percent of women of reproductive age (Shafrir et al., 2018) and is associated with severe dysmenorrhoea, chronic pelvic pain, dyspareunia, bowel and bladder symptoms, fatigue and infertility, with substantial negative impacts on quality of life.

The aetiology of endometriosis has been widely debated. Retrograde menstruation, defined as the reflux of menstrual blood and viable endometrial cells into the peritoneal cavity, has long been proposed as a central mechanism of disease development (Sampson, 1927). Yet despite occurring in approximately ninety percent of menstruating individuals, only a minority develop persistent endometriotic lesions (Mehedintu et al., 2014). Additional factors, including altered innate immune responses, impaired clearance of ectopic cells, local oestrogen production and a heightened inflammatory environment, are thought to facilitate ectopic lesion establishment and persistence within the peritoneal environment (Zondervan et al., 2018).

Altered iron handling has emerged as an important component of the peritoneal microenvironment in endometriosis. Retrograde menstruation introduces red blood cells into the peritoneal cavity, where their breakdown releases haem and free iron (Ng et al., 2020). This can lead to local iron, ferritin and haemosiderin accumulation within lesions and surrounding tissues and immune cells such as iron-laden macrophages. Excess iron promotes oxidative stress and inflammation, and may impair ferroptosis, an iron-dependent form of regulated cell death, potentially allowing ectopic endometrial-like cells to evade iron-mediated cell death and persist (Ng et al., 2020).

These processes may be particularly accentuated in more extensive disease, which also has been associated with higher heritability (Rahmioglu et al., 2023). For example, endometriomas exhibit iron-rich microenvironments dominated by macrophages containing heme and iron (Wölfler et al., 2013) and deep endometriosis has also been linked to oxidative stress pathways, through generation of superoxide anions, hydrogen peroxide, and nitric oxide (Tosti et al., 2015). Together, these findings reinforce that local iron accumulation contributes to the inflammatory and oxidative milieu characteristic of endometriosis.

In contrast to this local iron overload, epidemiological studies suggest that women with endometriosis may exhibit reduced systemic iron levels. Heavy menstrual bleeding and chronic inflammation may both contribute to reduced systemic iron availability, and a large longitudinal cohort study reported a 46% increased risk of iron deficiency among women with endometriosis (Gete et al., 2024). However, findings across studies are inconsistent, and may be confounded by menstrual blood loss (Munro et al., 2023) (Ekroos et al., 2024) and inflammatory processes (Li and Li, 2026). Observational studies are therefore limited in their ability to distinguish whether altered systemic iron status contributes to disease development or arises as a consequence of endometriosis.

Systemic iron homeostasis is tightly regulated through coordinated processes of absorption, transport, storage and recycling. Circulating markers capture complementary aspects of this system, including serum iron (circulating levels), ferritin (iron storage), total iron-binding capacity (TIBC; reflecting transferrin-mediated transport capacity), transferrin saturation (TSP; reflecting iron availability) and hepcidin, the central hormonal regulatory of iron homeostasis.

Large-scale genome-wide association studies (GWAS) have identified genetic variants influencing these traits, providing well-characterised ‘instruments’ for investigating the genetic influence of iron homeostasis on other phenotypes and traits. For binary traits, GWAS identify small changes in the DNA (Single nucleotide polymorphisms, SNPs) of patients with a confirmed disease and assess whether these SNPs are more or less common compared to the general population. Any undiagnosed disease in the general population will be diluted out by the very large population which does not have the disease. Many GWAS-associated SNPs lie outside protein-coding regions but may be involved in regulating when, where and to what extent genes are expressed, thereby providing clues to the biological pathways underlying disease. For continuous traits, GWAS assess whether genetic variants are associated with higher or lower levels of the trait.

Concerning iron traits, the largest GWAS meta-analysis identified 123 regions of the genome containing genetic variants associated with serum iron, ferritin, TSP and TIBC at genome-wide significance (Moksnes et al. 2022). In addition, a large meta-analysis of circulating hepcidin identified 16 genomic regions containing variants significantly associated with hepcidin levels (Allara et al., 2024). Together, these genetic associations across complementary biomarkers of iron physiology provide a framework for investigating whether the genetic determinants of iron homeostasis overlap with those of endometriosis.

In parallel, GWAS of endometriosis to date have identified 42 regions of the genome containing genetic variants robustly associated with disease rsik, with subsequent analyses implicating genes and biological pathways related to reproductive biology, immune regulation and vascular processes (Rahmioglu et al., 2023). However, the extent to which the genetic determinants of iron homeostasis overlap with those of endometriosis, and whether systemic iron biomarkers have a causal role in disease risk, remains unclear.

In this study, we investigated the genetic relationship between iron homeostasis and endometriosis using the largest available GWAS datasets and complementary genomic approaches. We assessed genome-wide genetic correlations between systemic iron biomarkers and endometriosis, including analyses by disease severity, identified shared loci using multi-trait GWAS, and evaluated potential causal relationships using bidirectional Mendelian randomisation. Through these analyses, we aimed to clarify whether systemic iron homeostasis is linked to endometriosis risk and to provide insight into the biological mechanisms underlying this association.

## MATERIALS AND METHODS

### Data resources and quality control

This study utilised the most up-to-date largest available GWAS meta-analysis results for endometriosis and iron biomarkers as summarised in Table 1. All GWAS included individuals of European ancestry.

**Table 1.** Summary of GWAS meta-analysis datasets used in our study.

| Phenotype | Sample Size | Sex | Reference |
| --- | --- | --- | --- |
| Overall Endometriosis | 28,281 cases: 506,494 controls | Female-only | *Rahmioglu et al. (2023)* |
| Stage III/IV Endometriosis | 9,073 cases: 506,494 controls | Female-only | *Rahmioglu et al. (2023)* |
| Serum Iron | 236,612 | Sex-combined | *Moksnes et al. (2022)* |
| Ferritin | 257,953 | Sex-combined | *Moksnes et al. (2022)* |
| TIBC | 208,422 | Sex-combined | *Moksnes et al. (2022)* |
| Transferrin Saturation | 198,516 | Sex-combined | *Moksnes et al. (2022)* |
| Hepcidin | 91,675 | Sex-combined | *Allara et al. (2024)* |

GWAS meta-analysis results were downloaded as GRCh37, restricted to HapMap3 SNPs and filtered for minor allele frequency (MAF) greater than 0.01, while also excluding palindromic variants. Allele orientation was harmonised to ensure consistency in effect direction across traits. Analyses were restricted to autosomal variants. In addition, the extended major histocompatibility complex (MHC) region on chromosome 6 (approximately 24–35 Mb) had been removed. The rationale for deleting this region is that it is characterised by highly complex linkage disequilibrium patterns, extensive haplotypic structure, and substantial genetic diversity, which complicate the interpretation of association signals. All GWAS results datasets were harmonised using the LDSC munge_sumstats pipeline (https://github.com/bulik/ldsc/blob/master/munge_sumstats.py).

### Genetic correlation analysis

Genetic correlation quantifies the extent to which genetic effects are shared between two traits: a positive genetic correlation indicates that genetic variants associated with higher levels of one trait tend to be associated with higher levels, or greater genetic liability, of the other, whereas a negative genetic correlation indicates that their genetic effects tend to act in opposite directions. Genome-wide genetic correlations between traits were estimated through linkage disequilibrium score regression (LDSC), using the LDSC (v1.0.1) on GitHub (Bulik-Sullivan and Finucane, 2019). Analyses utilised precomputed LD scores derived from European ancestry individuals from the 1000 Genomes Project Phase 3 reference panel. Pairwise correlations were calculated between each iron biomarker and overall endometriosis, as well as for stage III/IV disease separately. Genetic correlations were also estimated between each of the iron traits in order to determine how similar their genetic basis is.

A Benjamini-Hochberg FDR correction (Benjamini and Hochberg, 1995) was applied to account for multiple testing across the 10 pairwise genetic correlations estimated in the LDSC analysis, comprising five iron traits tested against overall endometriosis and the same five iron traits tested against stage III/IV endometriosis. An FDR-based approach was considered more appropriate than Bonferroni correction, due to the high level of correlation between the iron-related biomarkers

### Multi-trait analysis of GWAS

Multi-trait analysis of GWAS (MTAG) leverages shared genetic architecture between genetically correlated traits to increase power to identify associated variants, while generating trait-specific association estimates (Turley et al., 2018). In this study, MTAG was used to investigate whether leveraging shared genetic architecture between endometriosis and iron biomarkers could identify genetic associations that were not detected when each trait was analysed independently.

MTAG (v1.0.8; https://github.com/JonJala/mtag) was applied pairwise, jointly analysing endometriosis with one iron-related biomarker at a time rather than modelling all traits simultaneously. Summary statistics were harmonised using the MTAG-specific mtag_munge.py pipeline to standardise formatting, align alleles, remove strand-ambiguous variants, and restrict analyses to high-quality SNPs suitable for cross-trait analysis. Trait-specific association estimates were generated separately for endometriosis and each iron-related biomarker.

MTAG assumes a genome-wide homogeneous variance–covariance structure of SNP effects and uses LD score regression to estimate genetic covariance and estimation error, including that arising from sample overlap (Turley et al., 2018). The suitability of these assumptions was assessed in the context of the polygenic architecture of the traits and by inspection of LD score regression intercepts.

### Lead SNP identification and locus overlap

Independent genetic loci were defined using LD-based clumping. Genome-wide significant variants (P < 5 × 10⁻⁸) were evaluated sequentially, and a variant was retained as an independent lead SNP if no previously selected lead SNP was located within ±1 Mb and pairwise linkage disequilibrium was low (r² < 0.01).

This approach was first applied to the univariate endometriosis GWAS to define genome-wide significant loci. The same procedure was then used for the MTAG-derived results to assess whether joint analysis with iron biomarkers increased discovery of significant loci. Variants identified through MTAG were considered novel if they reached genome-wide significance and showed at least suggestive association (P < 5 × 10⁻⁵) in the univariate endometriosis GWAS.

To identify shared genetic loci between endometriosis and iron biomarkers, genome-wide significant lead SNPs from each analysis were compared. Overlap was defined as identical lead SNPs or lead SNPs located within ±1 Mb. Loci meeting the latter criterion were considered shared loci. To determine whether overlapping loci reflected the same underlying genetic signal, pairwise linkage disequilibrium (LD; r²) between lead SNPs was assessed using the LDlink LDpair tool (https://ldlink.nih.gov/). LD ≥ 0.2 was considered evidence that variants likely represent the same underlying association signal.

### Functional annotation using GTEx and Genomic Context

eQTLs and sQTLs are genetic variants associated with differences in the amount of gene expression and RNA splicing, respectively, with these regulatory associations can differ between tissues. We therefore used eQTL and sQTL data to investigate whether variants at the identified loci were associated with regulation of nearby or distant genes, and to identify candidate genes and tissues through which these genetic associations might act. Expression quantitative trait locus (eQTL) and splicing quantitative trait locus (sQTL) data were obtained from the Genotype-Tissue Expression (GTEx) Project (v8; https://gtexportal.org/home/) (The GTEx Consortium, 2020). Two sets of variants were evaluated: (i) novel genome-wide significant endometriosis loci identified through MTAG, and (ii) genome-wide significant loci shared between endometriosis and iron biomarkers.

Variants were queried across selected tissues relevant to iron metabolism and endometriosis, including liver, small intestine, spleen and whole blood, as well as reproductive tissues including uterus, ovary and fallopian tube. Whole blood eQTLGen data were additionally included to increase power for detecting regulatory associations. Genomic context was annotated using the Ensembl Variant Effect Predictor (VEP) (https://www.ensembl.org/info/docs/tools/vep/index.html) to classify variants according to their predicted functional consequence (e.g., intronic, intergenic, missense, UTR variants).

### Bidirectional Mendelian Randomisation

Bidirectional Mendelian randomisation (MR) uses genetic variants associated with an exposure as instrumental variables to investigate potential causal relationships between two traits in both directions. Here, two-sample MR was used to test whether genetically predicted levels of iron biomarkers were associated with endometriosis risk and, conversely, whether genetic liability to endometriosis was associated with levels of iron biomarkers. Analyses were conducted in R using the TwoSampleMR package (v0.6.30) (Hemani et al., 2020).

Genome-wide significant variants (p < 5 × 10⁻⁸) were selected as instrumental variables. To reduce correlation between instruments due to linkage disequilibrium (LD), variants were clumped within a 10,000 kb window at three LD thresholds ( r² = 0.1, 0.01, and 0.001), using the 1000 Genomes European reference panel and the ld_clump() function from the ieugwasr package. Analyses were repeated at each LD threshold to assess the robustness of findings to different levels of LD pruning.

The inverse-variance weighted (IVW) method was used as the primary MR analysis (Burgess, Butterworth and Thompson, 2013). Weighted median (Bowden et al., 2016) and MR-Egger regression (Jack Bowden, Davey Smith and Burgess, 2015) were performed as sensitivity analyses because they make different assumptions regarding potential horizontal pleiotropy, whereby genetic variants influence the outcome through pathways other than the exposure of interest. Evidence of directional horizontal pleiotropy was assessed using the MR-Egger intercept test (J. Bowden, Davey Smith and Burgess, 2015), and heterogeneity between variant-specific estiamtes was assessed using Cochran’s Q statistic (Bowden et al., 2019).

MR-PRESSO was additionally used to identify potential pleiotropic outlier variants and to reassess causal estimates following their removal (Verbanck et al., 2018). Instrument strength was evaluated using the F-statistic and proportion of variance explained, with F > 10 suggesting sufficient instrument strength. To account for multiple testing across five iron traits and three LD thresholds (15 tests), p-values were adjusted using the Benjamini–Hochberg false discovery rate (FDR) method (Benjamini and Hochberg, 1995).

Statistical power was assessed using the mRnd MR power calculator (https://shiny.cnsgenomics.com/mRnd/) (Brion, Shakhbazov and Visscher, 2013). For analyses with endometriosis as the binary outcome, power was estimated across hypothetical effect sizes corresponding to odds ratios (ORs) of 1.01, 1.05, 1.10, 1.15 and 1.20, together with the corresponding inverse effects. For analyses with iron biomarkers as continuous outcomes, power was estimated for effect sizes of β = 0.01, 0.05, 0.10, 0.15 and 0.20 and the corresponding negative effects, consistent with the parameterisation of the mRnd calculator.

### Pleiotropy assessment

Pleiotropy occurs when a genetic variant influences more than one trait or biological process. In Mendelian randomisation, horizontal pleiotropy occurs when an instrumental genetic variant influences the outcome through a pathway other than the exposure being investigated, potentially biasing the causal estimate. In the present analysis, horizontal pleiotropy was further assessed using MR-PRESSO to identify outlier variants and evaluate whether these variants distorted the causal estimates (https://github.com/rondolab/MR-PRESSO) (Ron Do Laboratory, 2025) (Verbanck et al., 2018). Analyses were performed using 5,000 simulations (NbDistribution = 5000) and a significance threshold of P < 0.05. Where outliers were detected, causal estimates were reassessed following their removal. This procedure was applied across all LD clumping thresholds and in both causal directions.

### Ethical approval

This study used only publicly available, fully anonymised GWAS summary statistics and did not involve new data collection. Ethical approval was therefore not required under the policies governing secondary analyses of de-identified genomic data. Analyses complied with all requirements for the use of publicly available genetic datasets and with the ethical principles outlined by Human Reproduction.

## RESULTS

### Genetic correlations indicate reduced systemic iron availability in endometriosis

We examined genetic correlations between iron biomarkers and endometriosis to assess shared genetic architecture. Strong correlations were observed between iron biomarkers themselves, consistent with established iron physiology. In particular, serum iron and TSP were strongly positively correlated (r_g_ = 0.73, p = 1.31 x 10^-5^), as were ferritin and hepcidin (r_g_ = 0.73, p = 4.78 x 10^-41^), while TIBC showed inverse correlations with ferritin (r_g_ = −0.29, p = 2.87 x 10^-9^) and TSP (r_g_ = −0.50, p = 8.24 x 10^-26^), reflecting its role as a marker of reduced iron availability. Genetic correlations between iron biomarkers and endometriosis showed a consistent pattern indicative of increased endometriosis liability associated with reduced systemic iron availability (Figure 1, Supplementary Table 1). Markers reflecting circulating iron availability were inversely correlated with genetic risk of endometriosis, including ferritin (r_g_ = −0.10, p = 0.022), TSP (r_g_ = −0.16, p = 0.006), and hepcidin (r_g_ = −0.13, p = 0.042), with a weaker inverse correlation observed for serum iron (r_g_ = −0.09, p = 0.10).

**Figure 1.**
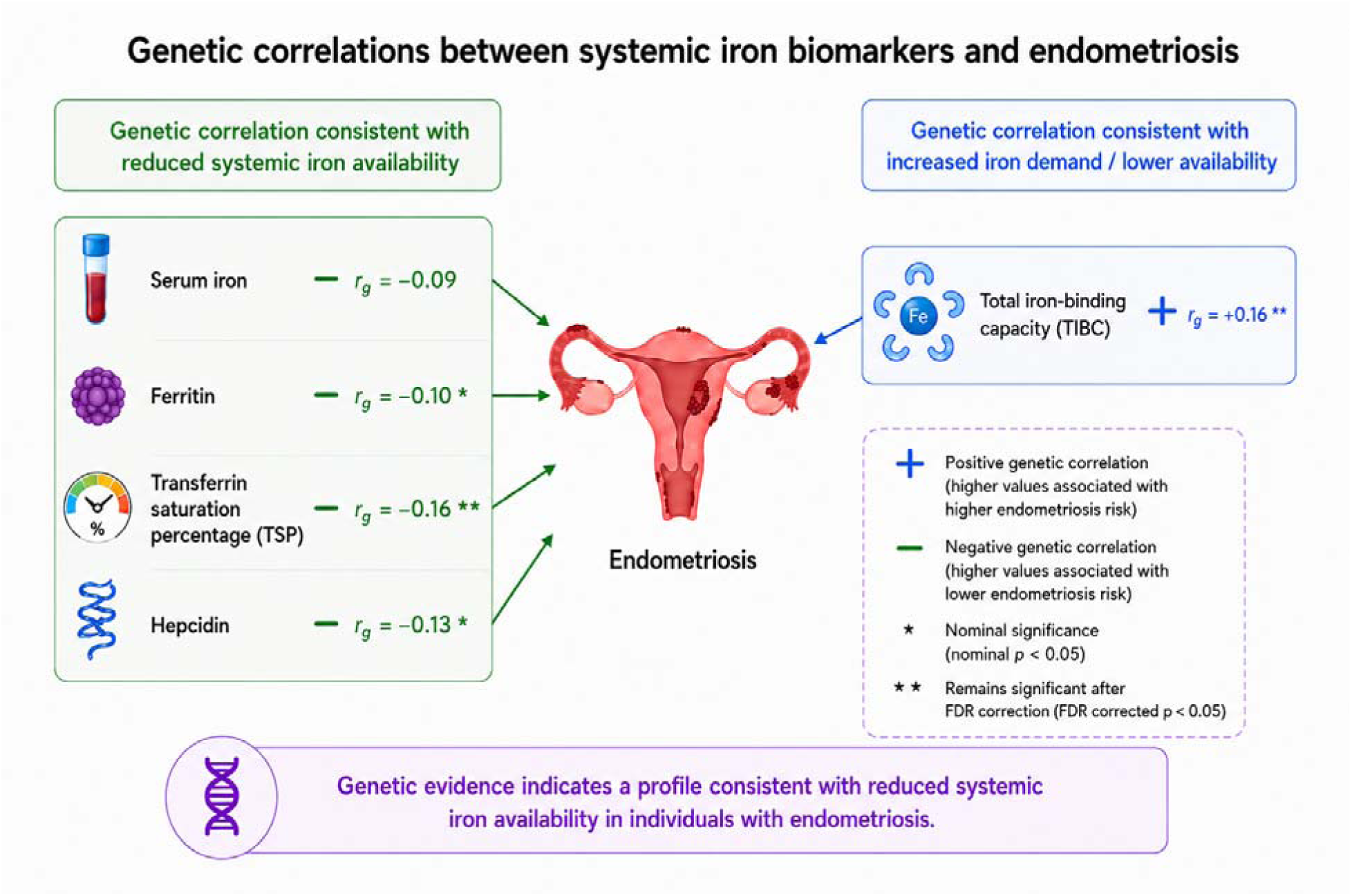
Genetic correlation results using LDSC between systemic iron biomarkers and overall endometriosis using GWAS summary statistics. Genetic correlation estimates for stage III/IV endometriosis, alongside LDSC regression diagnostics including SNP heritability estimates, standard errors, and intercepts for heritability and genetic covariance, are provided in Supplementary Table 1.

In contrast, TIBC demonstrated a positive genetic correlation (r_g_ = 0.16, p = 4 x 10_-4_). After FDR correction for multiple parallel testing, associations for TIBC (FDR-adjusted p = 0.004) and TSP (FDR-adjusted p = 0.031) remained statistically significant, whereas those for ferritin (FDR-adjusted p = 0.075) and hepcidin (FDR-adjusted p = 0.075) were attenuated. Together, these findings indicate that increased genetic risk of endometriosis is consistent with reduced systemic iron availability.

Analyses restricted to stage III/IV endometriosis showed directionally consistent results, although the statistical evidence for these associations was weaker (Figure 2, Supplementary Table 1). The magnitude and direction of genetic correlations for all serum iron (overall r_g_ = −0.088, FDR-adjusted p = 0.125; stage III/IV r_g_ = −0.143, FDR-adjusted p = 0.124), ferritin (overall r_g_ = −0.099, FDR-adjusted p = 0.075; stage III/IV r_g_ = −0.084, FDR-adjusted p = 0.178), TIBC (overall r_g_ = 0.159, FDR-adjusted p = 0.004; stage III/IV r_g_ = 0.128, FDR-adjusted p = 0.075), TSP (overall r_g_ = −0.158, FDR-adjusted p = 0.031; stage III/IV r_g_ = −0.157, FDR-adjusted p = 0.075) and hepcidin (overall r_g_ = −0.130, FDR-adjusted p = 0.075; stage III/IV r_g_ = −0.031, FDR-adjusted p = 0.764) remained broadly stable, supporting a consistent signal of reduced iron availability in more extensive disease. Although slight differences in effect size in stage III/IV disease with respect to overall endometriosis may reflect biological variation in systemic iron handling in advanced disease, estimates were imprecise and not statistically significant, likely due to reduced sample size and power in stage III/IV restricted GWAS (Figure 2, Supplementary Table 1). Given the greater statistical power of the full dataset, subsequent analyses focused on overall endometriosis.

**Figure 2.**
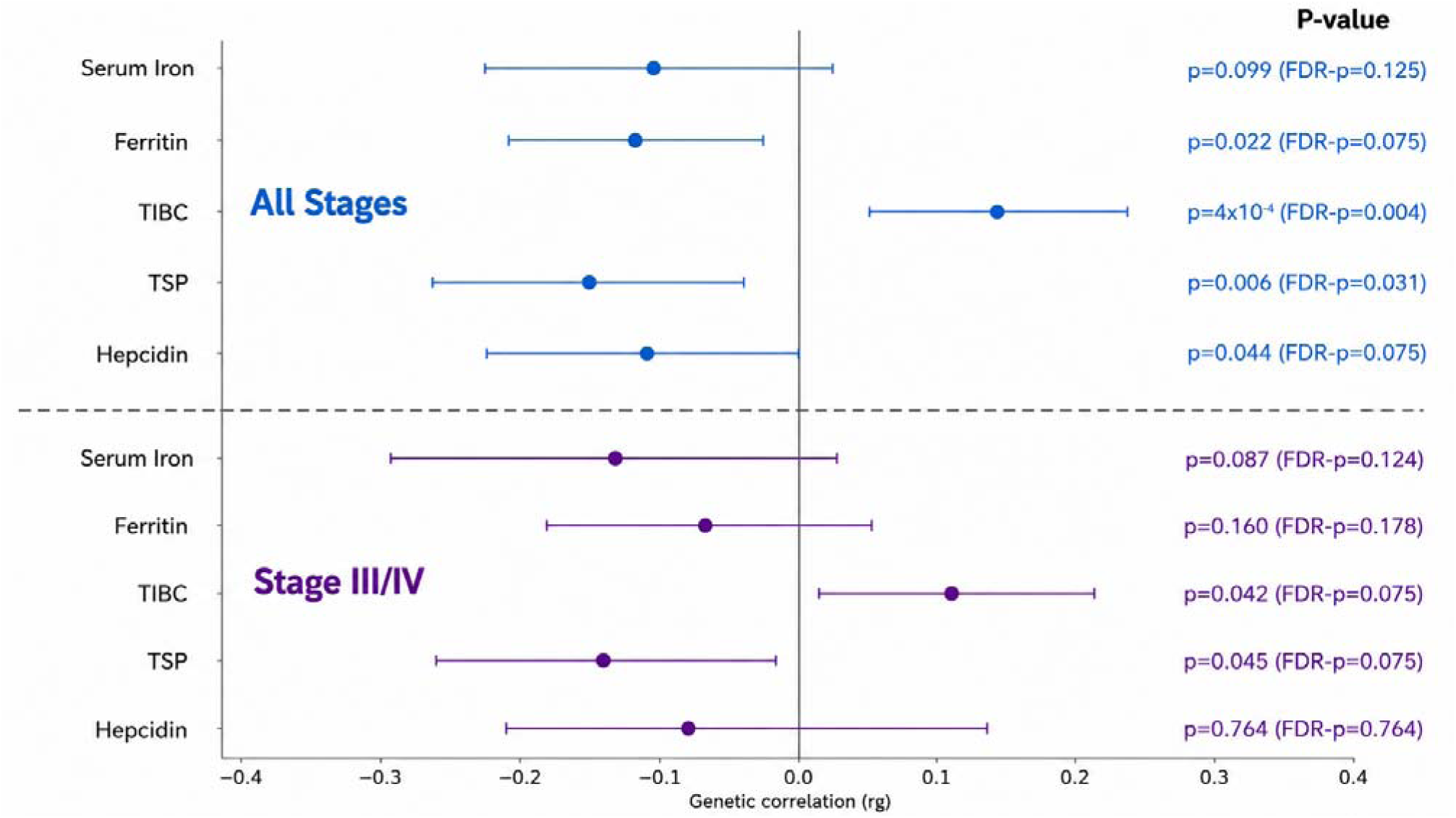
Genetic correlation results of overall and stage III/IV endometriosis with systemic iron biomarkers from LDSC using GWAS summary statistics. LDSC regression diagnostics, including SNP heritability estimates (h²), standard errors, and intercepts for heritability and genetic covariance, are provided in Supplementary Table 1.

### Leveraging shared genetic architecture with iron biomarkers enhances endometriosis locus discovery

We leveraged the shared genetic architecture between systemic iron biomarkers and endometriosis to improve locus discovery for endometriosis using MTAG. Across pairwise analyses, MTAG increased the number of genome-wide significant loci for endometriosis relative to the univariate GWAS (p < 5 × 10⁻⁸), identifying eight additional novel loci (Table 2). Full results and comparisons with univariate endometriosis GWAS are provided in Supplementary Tables 2-6.

**Table 2.**
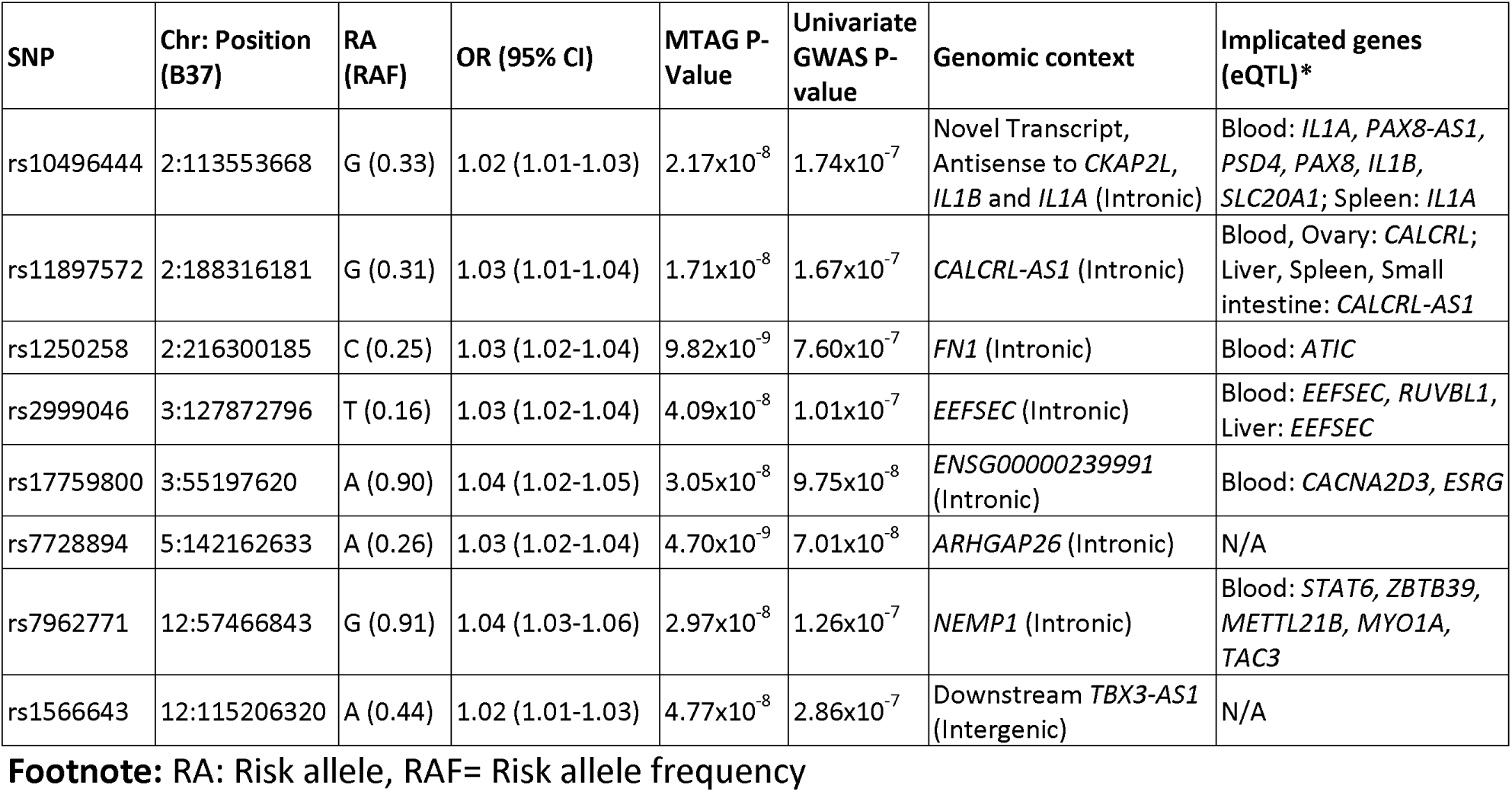
Novel 8 genome-wide significant endometriosis loci identified through MTAG with iron-related marker GWAS results. All genome-wide significant lead endometriosis variants from MTAG results across the 5 iron biomarkers are given in supplementary table 2-6.

Notably, loci at 2p13 (IL1A/IL1B) and 2q35 (FN1), previously implicated in endometriosis (Sapkota et al., 2017) but not reaching genome-wide significance in more recent GWAS (Rahmioglu et al., 2023), achieved genome-wide significance in MTAG analyses. Leveraging the shared genetic architecture between endometriosis and iron traits thereby strengthened evidence for the loci’s involvement in endometriosis risk.

Across all pairwise analyses, MTAG-derived association estimates were directionally consistent with univariate endometriosis effects but demonstrated modest shifts in effect size and precision, consistent with incorporation of cross-trait covariance. Notably, many of these additional loci were near genome-wide significance in the univariate endometriosis GWAS and surpassed the P < 5 × 10⁻⁸ threshold only after MTAG, indicating increased effective power revealing novel signals.

Functional annotation of the remaining loci further highlighted biologically relevant pathways. Variants near CALCRL suggest roles in vascular signalling (Selvarajan et al., 2024), while loci mapping to ARHGAP26 and EEFSEC implicate cellular signalling (Long et al., 2025) and protein synthesis pathways (Simonović and Puppala, 2018). Additional signals, including those near NEMP1 and TBX3-AS1, point towards regulatory mechanisms potentially involved in tissue remodelling (Tsatskis et al., 2020) and gene expression control (Jauregi-Miguel et al., 2025). Collectively, these findings indicate that leveraging shared genetic architecture with iron-related traits can enhance detection of biologically relevant endometriosis loci.

### Shared genetic loci link iron homeostasis and endometriosis risk

To further characterise the shared genetic architecture between systemic iron biomarkers and endometriosis, we examined overlap between genome-wide significant lead variants identified in MTAG analyses. Across analyses, 8 loci demonstrate overlap between endometriosis and at least one iron-related marker (Table 3), indicating shared genetic signals.

**Table 3.** Shared genome-wide significant loci between endometriosis and systemic iron biomarkers from MTAG results. e/sQTLs were identified in female reproductive tissues including uterus, ovary, fallopian tubes, liver, spleen, small intestine, blood. Please see supplementary table 7 for detailed look-up of eQTLs per variant per tissue.

| Traits | SNP | Chr:Pos | A1 (A1F) | OR (95% CI) | P-Value | LD (r<sup>2</sup>) | Genomic context | Implicated genes (e/sQTL) |
| --- | --- | --- | --- | --- | --- | --- | --- | --- |
| Endometriosis | rs6025 | 1:169519049 | C (0.98) | 1.09 (1.06-1.12) | 4.25x10<sup>-10</sup> | 1 | *F5* (Missense variant) | Blood: *ATP1B1*, *NME7*, *METTL18*, *SLC19A2*, *SELL* Spleen: *SCYL3* |
| Ferritin | rs6025 | 1:169519049 | C (0.98) | 0.86 (0.84-0.88) | 3.15x10<sup>-30</sup> |  |  |  |
| TIBC | rs6025 | 1:169519049 | C (0.97) | 1.12 (1.09-1.14) | 5.09x10<sup>-20</sup> |  |  |  |
| TSP | rs6025 | 1:169519049 | C (0.97) | 0.92 (0.90-0.95) | 5.48x10<sup>-11</sup> |  |  |  |
| Endometriosis | rs3765350 | 1:22447316 | G (0.22) | 1.05 (1.04-1.06) | 7.02x10<sup>-25</sup> | 0.005 | *WNT4* (Intronic) | Blood: *CDC42*, *LINC00339*, *USP48*, *HSPG2*, *CDC42-AS1* |
| Ferritin | rs12568930 | 1:22702231 | C (0.18) | 1.03 (1.02-1.04) | 4.40x10<sup>-13</sup> |  | Downstream *WNT4* (Intergenic) | N/A |
| Endometriosis | rs1250258 | 2:216300185 | C (0.25) | 1.03 (1.02-1.04) | 9.82x10<sup>-9</sup> | 1 | *FN1* (Intronic) | Blood: *ATIC* |
| Ferritin | rs1250258 | 2:216300185 | C (0.32) | 0.98 (0.97-0.98) | 2.22x10<sup>-12</sup> |  |  |  |
| Endometriosis | rs10020668 | 4:55996903 | G (0.73) | 1.05 (1.04-1.06) | 8.55x10<sup>-25</sup> | 0.001 | Upstream *KDR* (Intergenic) | Blood, Spleen: *SRD5A3*<br>Blood: *CLOCK* |
| Ferritin | rs3749474 | 4:56300685 | C (0.64) | 1.02 (1.01-1.02) | 1.20x10<sup>-8</sup> |  | *TMEM165* (Intronic) | Blood, Small Intestine, Spleen: *TMEM165*<br>Liver, Blood, Small intestine, Spleen: *SRD5A3*<br>Ovary, Blood, Small Intestine Spleen: *CLOCK* |
| Endometriosis | rs17053711 | 8:25311269 | G (0.72) | 1.03 (1.02-1.04) | 1.73x10<sup>-8</sup> | 0.001 | *KCTD9* (Intronic) | Blood: *GNRH1*, *DOCK5* |
| Ferritin | rs7009973 | 8:23374454 | G (0.34) | 1.02 (1.01-1.03) | 1.07x10<sup>-9</sup> |  | Intergenic | Blood: *SLC25A37*, *LOXL2*, *NKX3-1* |
| Endometriosis | rs579459 | 9:136154168 | C (0.21) | 1.03 (1.02-1.04) | 1.44x10<sup>-11</sup> | 1 | Intergenic | Liver, Spleen, Vagina, Blood: *ABO*<br>Blood: *SURF1*, *GBGT1*, *CACFD1* |
| Ferritin | rs651007 | 9:136153875 | C (0.84) | 1.05 (1.04-1.06) | 3.44x10<sup>-30</sup> |  | Intergenic | Liver, Spleen, Vagina, Blood: *ABO*<br>Blood: *SURF1*, *GBGT1*, *CACFD1* |
| Endometriosis | rs17119407 | 12:57452308 | T (0.91) | 1.04 (1.03-1.05) | 4.87x10<sup>-8</sup> | 0.001 | *NEMP1* (3' UTR) | Blood: *STAT6*, *ZBTB39*, *MYO1A*, *METTL21B*, *TAC3* |
| Hepcidin | rs4760355 | 12:57725197 | A (0.27) | 0.97 (0.96-0.98) | 3.91x10<sup>-9</sup> |  | *R3HDM2* (Intronic) | Blood: *STAT6*, *METTL21B*, *TMEM194A*, *MARS*, *DDIT3*, *ATP23* |
| Endometriosis | rs749647 | 14:93080703 | C (0.65) | 1.02 (1.02-1.03) | 4.49x10<sup>-8</sup> | 0.001 | *RIN3* (Intronic) | N/A |
| Serum Iron | rs7151526 | 14:94863636 | C (0.96) | 0.95 (0.93-0.97) | 2.89x10<sup>-8</sup> |  | *SERPINA9* (Intronic) | N/A |

Three loci showed genome-wide significant association in both endometriosis and iron-related traits analyses with lead variants in perfect LD (r^2^=1), suggesting shared underlying causal variants (Table 3). The most consistent signal was observed at rs6025 on chromosome 1 (1q24.2), which was associated with endometriosis as well as multiple iron-related traits, including ferritin, TIBC and TSP. This variant corresponds to the Factor V Leiden mutation (F5), a well-characterised exonic missense variant, resulting in an amino acid substitution in the coagulation factor V protein (Sinclair and Poon, 2013). This locus highlights a potential link between iron homeostasis and coagulation pathways that may contribute to the inflammatory and vascular microenvironment implicated in endometriosis pathophysiology.

Additional shared loci likely tagging the same causal variants are observed at 2q35 (FN1) and 9q34.2 (ABO). The rs1250258 locus implicates FN1, a gene involved in extracellular matrix organisation and tissue remodelling (Soubeyrand et al., 2022), processes relevant to endometriotic lesion establishment and persistence (Garcia Garcia et al., 2022). The rs579459/rs651007 locus implicates ABO, which influences coagulation and inflammatory pathways (Johansson et al., 2015), suggesting a role for haematological and vascular mechanisms linking iron traits and endometriosis risk.

The remaining overlapping loci containing genome-wide significant association for both endometriosis and iron biomarkers presented with low pairwise LD, suggesting independent signals within the same genomic region (Table 3). The WNT4 locus implicates genes involved in reproductive development (Pitzer et al., 2021) and cell migration (Li et al., 2025) (CDC42, HSPG2), while the locus near KCTD9 highlights genes related to neuroendocrine signalling (Stevenson et al., 2012) and cellular motility (Frank et al., 2017) (GNRH1, DOCK5). At the chromosome 8 locus, signals implicate SLC25A37, a key mitochondrial iron transporter (Chen et al., 2009), alongside genes involved in extracellular matrix remodelling (LOXL2) (Peng et al., 2025). Loci near KDR and SRD5A3 suggest roles in angiogenesis (Krikun, 2012), circadian regulation (Bolsius et al., 2021) and steroid metabolism (Son et al., 2023), while the TMEM165 locus points to broader metal ion homeostasis (Foulquier et al., 2012). The chromosome 12 region implicates STAT6 and related genes, supporting involvement of immune and inflammatory pathways (Wang, Wang and Zha, 2021), alongside genes linked to cellular stress responses (Jauhiainen et al., 2012). Additional loci, including those near RIN3 and SERPINA9, suggest roles in vesicular trafficking (Shen et al., 2020) and immune function (Tang et al., 2013). Collectively, these findings highlight overlap between iron homeostasis and endometriosis across pathways including coagulation, angiogenesis, immune regulation and extracellular matrix remodelling.

### Bidirectional Mendelian randomisation suggests limited evidence for causal effects between iron status and endometriosis

To determine whether the observed genetic correlations and shared genetic architecture between endometriosis and iron biomarkers were indicative of causal relationships, we next performed bidirectional Mendelian randomisation (MR). MR uses genetic variants associated with an exposure as proxies for that exposure to investigate its potential causal effect on an outcome. Because genetic variants are determined at conception and generally precede the development of disease, this approach can reduce the influence of reverse causation and many environmental or behavioural confounders that can affect conventional observational associations. Genome-wide significant variants across multiple LD clumping thresholds (r² = 0.1, 0.01, 0.001) were used as instrumental variables (IVs) (Supplementary Table 8). IVs therefore represent genetic proxies for the exposure, with their associations with the exposure and outcome used to estimate the potential causal effect between the two traits. LD clumping limits correlation between IVs, with increasingly stringent r² thresholds selecting progressively more independent variants and thereby allowing the robustness of estimates to instrument selection to be assessed. Primary estimates were obtained using the inverse-variance weighted (IVW) analyses, with weighted median and MR-Egger regression used as sensitivity analyses. Overall, there was limited and inconsistent evidence supporting causal effects in either direction (Figure 2).

In forward MR, where variability in iron biomarkers was the exposure and endometriosis the outcome, a suggestive association was observed between genetically predicted ferritin levels and reduced endometriosis risk in IVW model (OR = 0.84, 95% CI 0.73–0.98, nominal p = 0.023, FDR-adjusted p = 0.115 at r² = 0.001), with directionally consistent estimates across clumping thresholds (Supplementary Table 8). However, this association was not supported by weighted median or MR-Egger analyses, indicating limited robustness (Figure 2). A nominal association was also observed for TSP at the most permissive threshold (nominal p = 0.021, FDR-adjusted p = 0.115, r² = 0.1), but this was not consistent across models. No evidence of causal effects was observed for serum iron, TIBC or hepcidin.

In reverse MR, where endometriosis was considered as the exposure and variability in iron-related traits as outcomes, genetic liability to endometriosis showed a more consistent suggestive association with increased TIBC across clumping thresholds (OR = 1.03, 95% CI 1.01– 1.05, nominal p = 0.009, FDR-adjusted p = 0.045 at r² = 0.001), although not all sensitivity analyses reached statistical significance. No consistent associations were observed for serum iron, TSP or hepcidin. A weak inverse association with ferritin was observed but was not consistent across methods (Figure 2, Supplementary Table 8).

Sensitivity analyses indicated substantial heterogeneity across instruments for several traits. However, MR-Egger intercept tests did not indicate strong evidence of directional pleiotropy. MR-PRESSO identified a small number of outlier variants Supplementary Table 9a), but their removal did not materially alter the results (Supplementary Table 9b and 9c).

**Figure 2.**
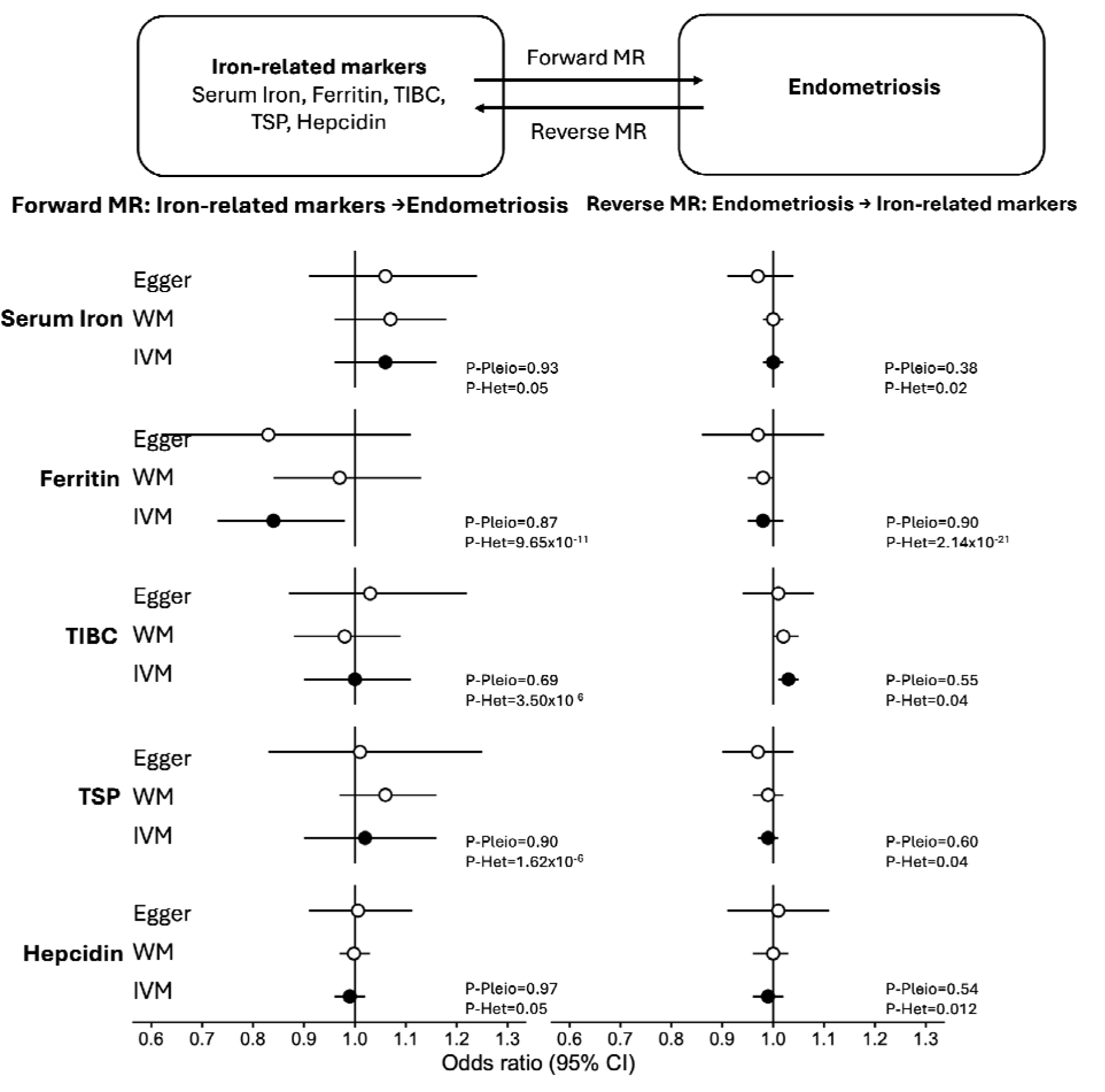
Bi-directional mendelian randomisation analysis between endometriosis (IVs=32) and iron biomarkers (Serum Iron IVs=22, Ferritin IVs=54, TIBC IVs=43, TSP IVs=22, Hepcidin IVs=32). IVs were defined using an LD clumping threshold of r =0.001. Filled circles denote main model inverse-variance weighted (IVM) estimates; open circles denote weighted median (WM) and MR-Egger estimates. P-values displayed in the figure are nominal p-values; FDR-adjusted p-values and full MR results across all linkage disequilibrium thresholds are provided in Supplementary Table 8.

Instrument strength exceeded conventional thresholds across all analyses (F > 10), indicating low risk of weak instrument bias. However, the proportion of variance explained by the genetic instruments was modest for several traits, particularly ferritin (R² = 0.023 – 0.038), serum iron (R² = 0.028 – 0.080) and hepcidin R² = 0.010–0.015) while for TIBC (R² = 0.043 – 0.188) and TSP (R² = 0.035 – 0.153) showed relatively higher variance explained. Power calculations indicated that the analyses were well powered to detect moderate effect sizes (OR ≥ 1.1–1.5) but had limited power to detect small effects (OR = 1.01–1.10) (Supplementary Table 10). Accordingly, the absence of consistent causal effects should be interpreted with caution, as small but potentially meaningful effects may not have been detectable.

## DISCUSSION

In this study, we provide genetic evidence linking systemic iron homeostasis to endometriosis. Across complementary analyses, we observed a consistent pattern indicating that genetic liability to endometriosis is associated with reduced systemic iron availability, supported by significant positive genetic correlation with TIBC (r_g_= 0.16, nominal p = 0.0004, FDR-adjusted p = 0.004) and negative correlation with TSP (r_g_ = −0.16, nominal p = 0.006, FDR-adjusted p = 0.031) and serum ferritin (r_g_= −0.10, nominal p = 0.022, FDR-adjusted p = 0.075). In addition, we identified eight genomic regions containing genome-wide significant associations with both endometriosis and iron homeostasis. These findings extend existing knowledge of iron dysregulation in endometriosis, which has largely focused on local iron accumulation within endometriotic lesions and the peritoneal environment (Wyatt et al., 2023), by demonstrating that the genetic architecture of endometriosis also overlaps with the underlying systemic iron homeostasis.

Reduced systemic iron availability and local iron accumulation are not necessarily contradictory, as iron status may differ substantially between the systemic circulation and local tissue environments. The apparent paradox of local iron excess within lesions alongside a genetic profile consistent with reduced systemic iron availability raises the possibility of compartmentalised dysregulation of iron handling, in which local iron-rich environments coexist with reduced circulating iron availability. Whether these processes are mechanistically connected, for example through altered iron trafficking or sequestration, or represent distinct manifestations of shared underlying biological processes remains to be established.

Notably, the observed genetic correlations, characterised by higher TIBC and lower ferritin and transferrin saturation, indicate that this systemic iron profile is captured at the level of germline genetic variation and is therefore independent of confounding by menstrual blood loss (Munro et al., 2023) (Ekroos et al., 2024) or secondary inflammatory processes (Li and Li, 2026) that complicate observational studies (Gete et al., 2024). This suggests that altered systemic iron handling may represent an intrinsic feature of endometriosis susceptibility rather than solely a downstream consequence of disease. Shared loci implicating coagulation, angiogenesis, extracellular matrix remodelling and immune and vascular pathways further support the involvement of systemic processes linking iron homeostasis and endometriosis. While Mendelian randomisation analyses provided only suggestive and inconsistent evidence for causal effects, the overall concordance across analytical approaches support that inherited susceptibility to endometriosis and systemic iron homeostasis share biological determinants, rather than reduced systemic iron status necessarily being solely a downstream consequence of disease.

At the locus level, three regions showed genome-wide significant associations with both endometriosis and iron biomarkers, with lead variants in perfect linkage disequilibrium (r² = 1), providing strong evidence that the associations arise from the same underlying genetic regions. The most striking signal was rs6025, the Factor V Leiden missense variant in F5, which was associated with endometriosis as well as ferritin, TIBC and TSP. F5 directly implicates coagulation and haemostatic processes (Tinholt et al., 2014), providing a potential biological connection between endometriosis risk and systemic iron balance through pathways influencing bleeding and iron loss. eQTL signals at this locus further implicated genes including SELL, involved in leukocyte adhesion and trafficking (Ivetic, Hoskins Green and Hart, 2019), and linking immune cell recruitment to inflammatory processes known to regulate iron sequestration, and SLC19A2, a thiamine transporter essential for erythroid function, providing a connection to red blood cell metabolism and iron utilisation (Diaz et al., 1999). At rs1250258, the involvement of FN1 points to extracellular matrix remodelling (Soubeyrand et al., 2022), a key process in endometriotic lesion establishment and fibrosis (Garcia Garcia et al., 2022), which may also shape local hypoxic and inflammatory environments that influence iron handling and oxidative stress, while ATIC supports cellular proliferation and metabolic activity (Li et al., 2017). At the chromosome 9 locus (rs579459/rs651007), eQTL evidence prioritises ABO, a gene with well-established effects on coagulation factors and systemic inflammation (Johansson et al., 2015), reinforcing the role of haematological regulation in both traits. Collectively, these loci converge on interconnected pathways involving haemostasis, immune activation, erythroid metabolism, and extracellular matrix remodelling, supporting a model in which variation in bleeding dynamics, inflammatory responses, and tissue repair processes jointly influence susceptibility to endometriosis and systemic iron homeostasis.

In contrast, 5 loci contained genome-wide significant associations for both endometriosis and iron-related traits but were represented by independent lead variants (r² < 0.01), suggesting potential convergence on shared biological pathways rather than a single shared causal variant. At the WNT4 locus, the endometriosis-associated variant implicates WNT4 alongside eQTL genes such as CDC42 and HSPG2, highlighting roles in reproductive tract development (Pitzer et al., 2021), cell migration (Prunskaite-Hyyryläinen et al., 2016) (Cohen et al., 2017), and extracellular matrix organization (Farach-Carson et al., 2014), while the corresponding iron-associated signal lacks clear functional annotation, suggesting partially distinct but potentially related mechanisms within this region. Similarly, at the chromosome 8 locus, iron-associated variation implicates SLC25A37, a key mitochondrial iron importer essential for heme biosynthesis (Chen et al., 2009), alongside LOXL2, involved in extracellular matrix remodelling and fibrosis (Peng et al., 2025), whereas the endometriosis signal (rs17053711) implicates genes such as DOCK5 and GNRH1, linking cell migration (Frank et al., 2017) and neuroendocrine regulation (Stevenson et al., 2012) to disease susceptibility. At the chromosome 4 locus, both traits converge on genes including CLOCK and SRD5A3, pointing toward shared influences of circadian rhythm (Bolsius et al., 2021) and steroid metabolism (Son et al., 2023), while KDR further supports a role for angiogenesis in endometriosis (Krikun, 2012) (Steinthorsdottir et al., 2016) and potentially iron distribution through vascular processes. Immune-related mechanisms are highlighted at the chromosome 12 locus, where both signals implicate STAT6, a key regulator of type 2 immune responses (Zhang et al., 2026), alongside genes involved in cellular stress (Jauhiainen et al., 2012) and metabolic regulation (Son et al., 2023) (DDIT3, MARS), supporting a link between immune activation, inflammation, and iron homeostasis. Additional loci, including those implicating TMEM165, RIN3, and SERPINA9, suggest roles in metal ion homeostasis (Foulquier et al., 2012), vesicular trafficking (Shen et al., 2020), and immune function (Tang et al., 2013), although their contributions are less well defined. Together, these findings suggest that shared genetic architecture between endometriosis and iron homeostasis is distributed across multiple interconnected biological systems rather than driven by a single mechanism.

Leveraging cross-trait genetic covariance through MTAG modestly increased locus discovery for endometriosis, identifying eight novel endometriosis loci beyond previously published GWAS meta-analysis (Rahmioglu et al., 2023). Importantly, all novel loci were near genome-wide significance in the univariate analysis, indicating that MTAG increased power to detect borderline associations rather than producing genome-wide significant findings in the absence of univariate support. Notably, two of these were previously reported associated with endometriosis (Sapkota et al., 2017) were not replicated at a genome-wide level in the latest published endometriosis GWAS meta-analysis (Rahmioglu et al., 2023), namely, 2p13 (IL1A/IL1B) and 2q35 (FN1). At 2p13 (IL1A/IL1B), the locus encompasses key pro-inflammatory cytokines IL-1α and IL-1β, which are central mediators of innate immune responses (Dinarello, 2018). Both cytokines are elevated in the peritoneal fluid of women with endometriosis (Kondera-Anasz et al., 2005) (Akoum et al., 2008) and contribute to a pro-inflammatory microenvironment that promotes lesion establishment, angiogenesis, and pain sensitisation (Machairiotis, Vasilakaki and Thomakos, 2021). IL-1 signalling can also stimulate the production of prostaglandins (Byron et al., 2023) and matrix-degrading enzymes (Fan et al., 2007), further supporting tissue invasion and remodelling (Machairiotis, Vasilakaki and Thomakos, 2021). Importantly, inflammatory cytokines such as IL-1 are known to influence systemic iron homeostasis through regulation of hepcidin and iron sequestration, linking chronic inflammation with altered iron metabolism (Inamura et al., 2005). This locus therefore highlights the importance of immune dysregulation as a shared mechanism between endometriosis and iron-related traits.

In addition, six novel endometriosis-associated regions were identified that were not reported before. The positional and eQTL evidence at these loci implicated genes involved in vascular, immune, and cellular regulatory processes. Notably, STAT6 (12q13.3) emerges as a compelling candidate, with experimental evidence supporting its role in endometriosis progression through promotion of inflammation and cell proliferation, and showing that inhibition of STAT6 signalling reduces lesion development in vivo (Lin et al., 2019). Similarly, CALCRL-AS1 (2q32.1) implicates CALCRL signalling with previous experimental evidence showing that CGRP/CALCRL pathways promote lesion development and fibrogenesis (Yan, Liu and Guo, 2019). The ARHGAP26 (5q31.3) locus supports roles in cytoskeletal regulation (Long et al., 2025) and cell migration (Chen et al., 2019), key features of endometriotic lesion invasion. Other implicated loci suggest roles for oxidative stress (EEFSEC, 3q21.3) (Simonović and Puppala, 2018), developmental regulation (TBX3-AS1, 12q24.21) (Khan et al., 2020) and calcium signalling (CACNA2D3, 3p21.1-p14.3) (Bellessort et al., 2018), whereas additional genes likely reflect broader regulatory processes.

Despite consistent genetic correlation and locus overlap, Mendelian randomisation did not provide robust evidence for causal effects between systemic iron biomarkers and endometriosis. Mendelian randomisation uses genetic variants as proxies for an exposure to test whether genetically predicted differences in that exposure are associated with an outcome providing evidence about potential causal relationships that is less susceptible to reverse causation and some forms of confounding than conventional observational analyses. In the forward direction, genetically predicted ferritin showed a suggestive inverse association at r² = 0.1 (OR=0.85, 95% CI=0.76–0.94; nominal-p = 0.002, FDR-adjusted p = 0.030) in the variance-weighted models, but estimates were attenuated with higher clumping thresholds, inconsistent across models. In the reverse direction, endometriosis liability was modestly associated with higher TIBC, consistent across clumping thresholds (r² = 0.001: OR=1.03, 95% CI=1.01–1.05, nominal-p = 0.009, FDR-adjusted p = 0.045; r² = 0.01: OR=1.03, 95% CI=1.01–1.05, nominal-p = 0.004, FDR-adjusted p = 0.045; r² = 0.1: OR=1.02, 95% CI=1.01–1.04, nominal-p = 0.006, FDR-adjusted p = 0.045) but findings were null in the weighted median model (Figure 2, Supplementary Table 8). Overall, the MR findings therefore do not establish that altered systemic iron status causes endometriosis, or that endometriosis liability directly causes altered systemic iron biomarkers.

Several factors may have limited the ability of the MR analyses to detect causal effects. Heterogeneity was common, particularly for ferritin and TIBC. For example, ferritin at r² = 0.1 demonstrated substantial heterogeneity (RSSobs = 215.826, P global < 2 × 10⁻⁴). MR-PRESSO identified outliers (rs1250258, rs6025, rs651007), but distortion tests were generally non-significant, and removal of outliers did not materially alter effect estimates (Supplementary Table 9). MR-Egger intercepts were consistently non-significant, providing little evidence of directional horizontal pleiotropy. Instrument strength represents a limitation. Although F-statistics exceeded conventional thresholds (F > 10), R² values were modest, particularly for ferritin (r²=0.001, R²=0.023; r²=0.01, R²=0.026; r²=0.1, R²=0.038) and hepcidin (r²=0.001, R²=0.010; r²=0.01, R²=0.011; r²=0.1, R²=0.015) (Supplementary Table 10). Such limited explanatory power reduces sensitivity to detect small causal effects, especially given the polygenic architecture of these traits. Moreover, variance explained ranged from roughly 1% for hepcidin to 19% for TIBC, further underscoring limited power to detect real effects.

Several additional limitations should be considered. The iron biomarker GWAS included both females and males, despite known sexual dimorphism in iron metabolism (Tao et al., 2023). Female-specific GWAS may therefore identify genetic effects that are particularly relevant to iron physiology in women but are attenuated in sex-combined analyses. Both the iron and endometriosis datasets were predominantly of European ancestry, limiting generalisability to other ancestral populations. Power was also substantially lower for stage III/IV endometriosis than for overall disease, restricting our ability to determine whether the genetic relationships with iron differ according to disease severity or phenotype. Functional annotation using eQTL and sQTL data is dependent on the tissues and sample sizes represented in available reference datasets, with relatively limited power in some female reproductive tissues. Furthermore, overlap between genome-wide significant signals does not by itself establish that the same causal variant or gene underlies associations with both traits; formal colocalisation, fine-mapping and functional validation will be required to resolve these relationships. Finally, the present genetic analyses cannot determine whether systemic and local iron dysregulation are mechanistically connected.

From a biological standpoint, our findings suggest that endometriosis genetic liability aligns with systemic iron restriction rather than systemic iron excess. This does not contradict the established accumulation of iron within endometriotic lesions and the peritoneal environment, but instead raises the possibility that local iron-rich microenvironments coexist with a genetically influenced systemic profile of reduced iron availability. Shared genetic pathways involving immune activation, coagulation, vascular function and tissue remodelling provide potential mechanisms through which both iron regulation and susceptibility to endometriosis may be influenced. Whether the systemic and local iron phenotypes represent mechanistically linked aspects of iron dysregulation or distinct consequences of shared biological pathways remains unknown.

From a clinical perspective, these findings suggest that reduced systemic iron availability may be a feature of the genetic architecture of endometriosis. They also indicate that associations between endometriosis and reduced systemic iron status should not necessarily be attributed solely to established factors such as menstrual blood loss or dietary iron intake. However, given the lack of robust causal evidence, it remains unclear whether altered iron status contributes directly to disease development or reflects downstream effects of shared biological pathways. Further research is needed to determine whether iron status has clinical relevance for disease progression or management.

Future work should prioritise female-specific GWAS of systemic iron biomarkers to capture sex-specific effects, alongside larger subtype-specific analyses of endometriosis beyond stage III/IV. In particular, distinguishing between ovarian endometrioma, peritoneal disease and deep disease may help clarify whether iron-related pathways differentially influence lesion phenotypes. Expanding analyses to more diverse ancestral populations will also be important to improve generalisability and identify population-specific genetic effects that may not be captured in European ancestry datasets

In conclusion, our findings demonstrate a shared genetic architecture between endometriosis and systemic iron homeostasis, characterised by genetic correlations consistent with reduced circulating iron availability. This genetic component suggests that the lower systemic iron status observed in women with endometriosis may not be attributable solely to consequences such as heavy menstrual bleeding or inadequate dietary iron intake, but may also reflect underlying inherited susceptibility. Shared genomic regions implicate coagulation, immune, vascular and tissue-remodelling pathways, suggesting that the relationship reflects overlapping polygenic biology rather than a single causal mechanism. Although the causal basis of this relationship remains uncertain, understanding how inherited variation in iron homeostasis and these interconnected biological pathways contributes to endometriosis may provide new insights into disease mechanisms and clarify the clinical significance of systemic iron status in endometriosis.

## Author’s Roles

V.D. performed all analyses. V.D., N.R., S.M. interpreted results. V.D. and N.R. wrote the manuscript. N.R., S.M., H.D., C.M.B. and K.T.Z. provided conceptual guidance, supervision and critical revisions. S.M. served as the line manager of this fellowship at the Oxford Big Data Institute. All authors contributed to interpretation of results and approved the final manuscript.

## Data Availability

All data used in this study are publicly available. Endometriosis GWAS summary statistics (Rahmioglu et al., 2023) are available from the NHGRI-EBI GWAS Catalog under accession GCST90205183. Iron biomarker (serum iron, ferritin, TIBC, transferrin saturation) GWAS summary statistics (Moksnes et al., 2022) are available from NTNU Open Research Data. Hepcidin GWAS summary statistics (Allara et al., 2024) are available from the NHGRI-EBI GWAS Catalog under accession GCST90451683.

https://www.ebi.ac.uk/gwas/studies/GCST90205183

https://dataverse.no/dataset.xhtml?persistentId=10.18710/S9TJEL

https://www.ebi.ac.uk/gwas/publications/39643614

## Acknowledgements

Computation used the Oxford Biomedical Research Computing (BMRC) facility, a joint development between Centre for Human Genetics and the Big Data Institute supported by Health Data Research UK and the NIHR Oxford Biomedical Research Centre. The views expressed are those of the author(s) and not necessarily those of the NHS, the NIHR or the Department of Health.

